# Individual-level deviations from normative brain morphology in violence, psychosis, and psychopathy

**DOI:** 10.1101/2023.10.29.23297735

**Authors:** Unn K Haukvik, Thomas Wolfers, Natalia Tesli, Christina Bell, Gabriela Hjell, Thomas Fischer-Vieler, Nina Bang, Ingrid Melle, Ole A Andreassen, Kirsten Rasmussen, Ingrid Agartz, Lars T Westlye, Christine Friestad, Jaroslav Rokicki

## Abstract

**Background:** Neuroimaging research has shown brain morphological abnormalities associated with violence and psychosis, but individual differences are substantial and results not consistent across studies. Recently developed normative modeling of brain MRI-features provides a possibility to parse this heterogeneity by mapping inter-individual brain characteristics, which has not yet been explored in forensic psychiatry.

**Methods:** We explored brain heterogeneity in persons with a history of severe violence with or without a schizophrenia spectrum disorder (HoV, n=58), non-violent patients with schizophrenia spectrum disorders (SSD-NV, n=138), and healthy non-violent controls (HC) from lifetime normative trajectories of cortical thickness, surface area, and subcortical volumes. Normative models were based on Freesurfer derived regions of interest from 58,836 individuals (ages 2-100) from 82 sites. Group differences and associations between brain deviations and psychopathy traits (PCL-R scores) were investigated.

**Results:** Across groups, we found an overall heterogeneous pattern of individual-level deviations, with a significantly higher frequency of extreme negative deviations in HoV (*p*=.020, *d*=.31) and SSD-NV (*p*=.019, *d=*.*48)*, than HC. Group differences were mostly present in subcortical volumes and cortical area, but not thickness, with significant regional group-level differences within the subcallosal and insular cortices, and the cerebellum, and no significant associations to psychopathy traits.

**Conclusion:** By applying normative modeling, this proof-of-concept study demonstrates the heterogeneous pattern of brain morphometry deviations associated with violence and psychosis. While the results warrant replication, studies addressing individual brain deviations may contribute to improved understanding of the complex underpinnings of violence in forensic psychiatry.

## Background

Violence committed by persons with severe mental disorders is a tragedy for the people involved, the treating health services, and the society at large. A key to prevent such events is to understand the complex biopsychosocial underpinnings of violence associated with severe mental disorders, their interaction with each other (Mitjans et al., 2019) and the situational characteristics of each violent act (Camus, Dan Glauser, Gholamrezaee, Gasser, & Moulin, 2021). This effort is challenged by the heterogeneity of disorders, symptom manifestations, substance use, personality traits and personal circumstances of the offender, as well as the multifaceted construct of violence, comprising instrumental, reactive, and psychosis driven actions (Stahl, 2014).

Over the last decades, neuroimaging research has tried to map the anatomy and functional characteristics associated with violence committed by persons with severe mental disorders (see (Fjellvang, Groning, & Haukvik, 2018) for review). Magnetic resonance imaging (MRI) studies have shown volumetric reductions of the orbitofrontal (Kumari et al., 2009) and anterior cingulate (Kumari et al., 2014) cortex, the amygdala and hippocampus (Del Bene et al., 2016), including their subfields and nuclei (Bell et al., 2022; Tesli et al., 2020), as well as cortical folding and thickness abnormalities (Kuroki et al., 2017; Narayan et al., 2007; Storvestre et al., 2019) in persons with psychosis and a history of severe violence. Aggression and impulsivity, which might be considered as proxies or precursors of violence, have been associated with similar volumetric reductions (Hoptman, Antonius, Mauro, Parker, & Javitt, 2014; Hoptman et al., 2005). This pattern of brain deviations aligns with regions and circuits involved in fear (Park & Chung, 2020), reward (Haber & Knutson, 2010), empathy (Engen & Singer, 2013), the processing of visual stimuli (Gerber, Golan, Knight, & Deouell, 2017), and the formation of positive psychosis symptoms including hallucinations (Cachia et al., 2020) and delusions (Tamminga, Stan, & Wagner, 2010). The findings vary across studies, which is expected considering the diversity of the violence and psychosis phenotypes (Schiffer et al., 2013). The traditional neuroimaging research statistical methodology comparing group averages does not account for this heterogeneity. As such, putative individual brain deviations and patterns pointing toward distinct phenotypic neurobiological underpinnings might remain undiscovered.

To supplement conventional group-based comparisons, statistical methods have been developed which model inter-individual deviations from the norm to address the heterogeneity of brain abnormalities associated with mental health disorders. The core concept of these models is based on pediatric growth charts, where individual variation is quantified against the growth information from a reference population (Rutherford et al., 2022). In mental health research, this approach has been translated into mapping inter-individual brain characteristics to reference maps of age-related brain anatomy, where mental health disorders have shown atypical trajectories of brain development and neurodegeneration (Bethlehem et al., 2022). As such, the application of normative models in neuroimaging research provides an alternative to the classical case-control or cluster analyses (Marquand et al., 2019). By using normative models, heterogeneous patterns of brain deviations have been mapped in schizophrenia and bipolar disorder, where patients showed large, but non-overlapping, individual deviations from the norm that complement the group mean differences (Wolfers et al., 2018). Large deviations from the norm have been demonstrated in antipsychotic naïve first episode psychosis patients (Remiszewski et al., 2022), and aggression has been linked to age-related brain deviations specifically in the amygdala in youth with disruptive behavior disorders (Holz et al., 2022). Normative modeling has also been used to map trait impulsivity and reward-related brain responses, and shown individual brain deviations to be associated with specific clinical symptoms (Marquand, Rezek, Buitelaar, & Beckmann, 2016).

Following recent advances in computational neuroscience, neuroimaging research has moved towards analyzing datasets from large publicly available databases or collaborative mega-analyses of data from individual research centers worldwide, which facilitates the disentanglement of neurobiological underpinnings of psychopathology on a large scale (Thompson et al., 2020). However, comorbidity between antisocial traits and psychosis leading to severe violence and violent offending occurs among a minority of persons and patients (Whiting, Gulati, Geddes, & Fazel, 2022). Hence, neuroimaging research in forensic psychiatry must aim at exploring methods that target the individual. Normative modeling provides a statistical approach which allows the detection of specific brain patterns associated with sub phenomena of the complex and heterogeneous construct of violence in severe mental disorders. The objective of this study is to assess the potential of normative modeling in forensic psychiatry neuroimaging research by investigating inter-individual patterns of deviations from the norm in persons with a history of severe violence with or without comorbid schizophrenia spectrum disorder (HoV), persons with a schizophrenia spectrum disorder but no history of violence (SSD-NV), and healthy controls with no history of violence (HC). We hypothesize that there will be high inter-individual variability for quantitative anatomic features of HoV compared with HC, in particular in regions previously associated with violence in severe mental disorders such as the amygdala, hippocampus, orbito-frontal cortex, and anterior cingulate cortex ((Fjellvang et al., 2018) for review). In line with previous literature (Wolfers et al., 2018), we expect deviations from the norm in SSD-NV, specifically in regions previously associated with schizophrenia, such as prefrontal cortices, hippocampus, thalamus, and the basal ganglia ((Haukvik, Hartberg, & Agartz, 2013; Haukvik, Tamnes, Soderman, & Agartz, 2018) for review). Moreover, we hypothesize that psychopathy traits will be associated with larger deviations (Johanson, Vaurio, Tiihonen, & Lahteenvuo, 2019).

## Methods

### Participants

Participants (*n*=782 male/608 female) were recruited from the Oslo area as part of four studies: the Thematically Organized Psychosis study (TOP) (Haukvik et al., 2015), the STROKEMRI study (Richard et al., 2020), The Youth-TOP study (uTOP) (Hilland et al., 2022), and The Forensic Psychiatry study (sTOP) (Bell et al., 2022; Tesli et al., 2022). 998 healthy controls (390 male/608 female) were used as a calibration set to adjust the model to our data (Rutherford et al., 2022). The remaining participants consisted of three groups: (1) persons with a history of severe violence (*HoV*, n=58), where 38 individuals had a comorbid schizophrenia spectrum disorder (SSD-V) and 20 individuals did not (nonSSD-V), (2) persons with no history of violence but with a schizophrenia spectrum disorder (*SSD-NV*, n=138), and (3) healthy controls (*HC*, n=196) matched by age and scanner. All were male due to the scarcity of eligible women within the participating forensic psychiatry units and prisons.

The participants had no history of head trauma that resulted in loss of consciousness, and no present or recent somatic disease that might have impacted brain morphology.

The *HoV* inclusion criteria were age between 18 and 70 years and a court or hospital record verified episode of severe violence, defined as homicide, attempted homicide, or physical (including sexual) violence towards other persons. The group comprised persons who were either sentenced or committed to compulsory mental health care in high security hospital wards or served a preventive detention sentence in a high security prison. The *SSD-NV* inclusion criteria required participants to be between 12 and 65 years old, with a DSM-IV diagnosis of schizophrenia spectrum disorder and no history of severe violence. The participants were included from hospital wards and out-patient clinics. *HC* were either drawn from the Norwegian national population registry (TOP, uTOP), or by newspaper ads and word of mouth (STROKEMRI). The HC inclusion criteria were age 12-18 (uTOP), 18-65 (TOP), or 20-88 (STROKEMRI) and no history of neurological or severe mental disorders.

The Norwegian Regional Committee for Medical Research Ethics approved the project which was conducted in accordance with the Helsinki declaration. All participants submitted written informed consent.

### Clinical assessment

Clinical characterization was performed by specially trained psychiatrists, medical doctors, and psychologists. Structured diagnostic assessment was performed with the SCID-I (M. B. First, Spitzer, R. L., Gibbon, M., Williams, J. B. W.,, 1996) (18 years or above), and the KIDDIE-SADS present and lifetime version (Kaufman et al., 1997) (below 18 years). The SCID II domains antisocial and borderline personality disorders were administered for HoV (M. B. First, Gibbon, M., Spitzer, R.L., Williams, J.B., Benjamin, L.S.,, 1997). The positive and negative symptom scale (PANSS) (Kay, Fiszbein, & Opler, 1987) was used to assess psychosis symptoms, the Wechsler Abbreviated Scale of Intelligence (WASI) for IQ (Wechsler, 2007), and substance use with AUDIT (Berman, Bergman, Palmstierna, & Schlyter, 2005) and DUDIT (Berman et al., 2005). Psychopathy traits were assessed with the Psychopathy Checklist Revised (PCL-R) supplemented by hospital records and court documents (Hare, 2003). The HC were assessed for mental disorders using the Primary Care Evaluation of Mental Disorders (Spitzer et al., 1994),

### MRI data acquisition

T1-weighted volumes were collected on two 3T scanners (GE Signa HDxt and GE DiscoveryTM-MR750) located at the Oslo University hospital, Norway. (1) GE Signa HDxt scanner with a standard 8-channel head coil, using a sagittal 3D fast spoiled gradient echo (FSPGR) sequence with the following parameters: repetition time (TR) = 7.8 ms, echo time (TE) = 2.9 ms, flip angle 12°, slice thickness 1.2 mm, 166 sagittal slices, field of view (FOV) 256×256 mm, acquisition matrix 256×192 mm, voxel size = 1×1×1.2 mm3 and (2) DiscoveryTM-MR750 scanner with the vendor’s 32-channel head coil, using an inversion recovery-fast spoiled gradient echo sequence (BRAVO) with the following parameters: TR = 8.16 ms, TE = 3.18 ms, TI = 450 ms, flip angle = 12°, FOV = 256 mm, acquisition matrix = 256×256 mm, 188 sagittal slices, slice thickness = 1.0 mm, voxel size = 1×1×1 mm^3^.

### Quality control and preprocessing

Image quality checking was carried out in two steps. First, all T1w images were processed using MRIqc (Esteban et al., 2017). Then two trained raters visually examined the scans marked for exclusion by MRIqc pipeline (NT and JR). The quality of the area and thickness of cortical maps was assessed in a comprehensive visual evaluation of lateral and medial snapshots of all maps. If the surface values had negative values, unusual patterns, or a significant value imbalance across hemispheres, the individual was removed. Freesurfer v7.1 was used to process data that passed quality control. Cortical thickness and area was summarized using the Destrieux atlas parcellation scheme (Destrieux, Fischl, Dale, & Halgren, 2010). Subcortical volume segmentation was based on a probabilistic atlas with priors for 40 brain structures (Fischl, 2012; Fischl et al., 2002). The ROIs used to train the initial normative models determined the selection of parcellation atlases for cortical and subcortical regions.

### Normative models

We used pretrained normative models from https://github.com/predictive-clinical-neuroscience/braincharts. Briefly, models were trained using data of 58835 individuals aged 2-100 from 82 sites. To model non-linear effects accurately, normative models were trained using Bayesian linear regression with likelihood warping using sinarcsinsh function (Fraza, Dinga, Beckmann, & Marquand, 2021). The dependent variable was set to the ROI values of volume, area, or thickness, while the independent variables were age and scanner. To summarize the cortical area and thickness, we used the regions described in a high-resolution Destrieux atlas (Destrieux et al., 2010). Models were both trained and applied to our data using python 3.8 and PCNtoolkit package (version 0.20). Further details on normative models are described in (Rutherford et al., 2022).To account for the absence of scanner data used to acquire data in the model, an additional harmonization step was implemented. An unmatched group of healthy controls (*n*=998) was utilized to fine-tune model parameters, enabling the application of normative models to unseen sites.

### Statistical analysis

To summarize inter-individual variation within each group (HoV, SSD-NV, and HC), we computed deviation scores (Z-scores) by separating them into positive and negative deviations. The percentage of subjects within each group with extreme positive (*Z*>2) and negative (*Z*<-2) deviations at a given ROI were compared and reported (Rutherford et al., 2022). Further, we examined which ROIs consistently exhibited extreme deviations in each modality (cortical area and thickness, subcortical volume) by quantifying the percentage of extreme deviations observed in each ROI for each diagnostic group. Then, we quantified the number of extreme deviations per individual to compare if participants from diagnostic groups had a higher count of extreme deviations in general as compared to HC. Additionally, we investigated the differences between SSD-NV and HoV groups. For completeness, all analyses were repeated with the HoV group stratified by comorbid schizophrenia status (SSD-V, nonSSD-V).

Subsequently, we conducted case-control group difference testing on the deviation scores. The R package matchIt was used to match age and scanning sites between HC and diagnostic groups using logistic regression distance with a 1:1 matching ratio (Ho, Imai, King, & Stuart, 2011). General linear models (GLM), as implemented in the permutation analysis of linear models (PALM) toolkit (Winkler, Ridgway, Webster, Smith, & Nichols, 2014), were used to perform the between-group statistical analyses, controlling for the effects of age and demeaning both data and independent variables in the design matrix. We used family wise error correction to account for multiple corrections: the number of ROIs, three modalities (cortical area and thickness, and subcortical volume) and 4 contrasts (Alberton, Nichols, Gamba, & Winkler, 2020). Additionally, we computed Cohen’s *d* effect sizes.

To determine whether there were group differences in the frequency of extreme positive and negative deviations, we performed permutation. The input data consisted of the number of extreme positive and negative deviations per individual, aggregated across all modalities. Specifically, we tested if the diagnostic groups had more negative deviations as compared to healthy controls and each-other, and if the opposite pattern was held for extreme positive deviations.

Finally, to examine the association between PCL-R scores and deviation scores in individuals with a history of violence, we used the GLM in the PALM toolbox with 10,000 permutations. The deviation scores of each ROI were treated as the dependent variable, while the PCL-R total score, age, psychosis status, and ICV (for area and volume) were included as independent variables.

## Results

### Demographics

Demographic characteristics are summarized in Table 1 (Supplementary Table 1 for HoV group stratified by psychosis status). Groups differed significantly on age, IQ score, and substance use (DUDIT), but not in alcohol use (AUDIT) and PANSS scores.

**Table 1.** Demographic and clinical characteristics.

|  | SSD-NV<br>(*n*=138) | HoV<br>(*n*=58) | HC<br>(*n*=196) | Total<br>(*n*=392) | P-value<br>*t* or *F*<br>test |
| --- | --- | --- | --- | --- | --- |
| Age (years) |  |  |  |  |  |
| Mean (SD) | 29.0 (8.69) | 37.4 (11.6) | 31.9 (9.99) | 31.7 (10.2) | *p*<0.001<br>*F*=15 |
| Median (min, max) | 26.9 (15.1, 57.8) | 36.7 (19.2, 71.0) | 29.3 (15.7, 70.9) | 29.1 (15.1, 71.0) |  |
| Wechsler abbreviated scale of intelligence (WASI-IQ) |  |  |  |  |  |
| Mean (SD) | 102 (14.4) | 97.5 (14.4) | 114 (11.0) | 107 (14.4) | *p*<0.001<br>*F*=41 |
| Missing | 10 (7.2%) | 22 (37.9%) | 44 (22.4%) | 76 (19.4%) |  |
| Positive and negative syndrome scale total (PANSS) |  |  |  |  |  |
| Mean (SD) | 60.8 (17.1) | 55.2 (20.4) | NA (NA) | 59.3 (18.2) | *p*=0.056<br>*t*=1.9 |
| Missing | 1 (0.7%) | 4 (6.9%) | 196 (100%) | 201 (51.3%) |  |
| Drug use disorders identification test (DUDIT) |  |  |  |  |  |
| Mean (SD) | 5.56 (8.88) | 9.47 (10.10) | 0.51 (1.63) | 3.67 (7.47) | *p*<0.001<br>*F*=30 |
| Missing | 14 (10.1%) | 28 (48.3%) | 67 (34.2%) | 109 (27.8%) |  |
| Alcohol use disorders identification test (AUDIT) |  |  |  |  |  |
| Mean (SD) | 6.60 (6.35) | 4.81 (4.59) | 6.05 (3.51) | 6.15 (5.05) | *p*=0.2<br>*F*=1.6 |
| Missing | 16 (11.6%) | 27 (46.6%) | 67 (34.2%) | 110 (28.1%) |  |

*Abbreviations:* SSD-NV - schizophrenia spectrum disorder patients without history of violence, HoV - participants with history of violence, HC - healthy controls, SD - standard deviation.

### Deviations from the normal trajectory

Extreme negative and positive deviations from the normal age related trajectory for each group are visualized in Figure 1 for cortical area, thickness, and subcortical volumes respectively (supplementary figures 1-3 present the same results with the HoV group stratified by psychosis status). On average, HC had extreme negative deviations in 1.51% of area, 0.95% of thickness, and 0.59% of subcortical volume regions. In contrast, SSD-NV participants had on average a 3.1% and HoV had 2.75% of extreme negative deviations in cortical area regions (Supplemental table 2 and 3). Further, HoV participants had more negative deviations in subcortical regions (2.42% when compared both with HC (0.59%) and with SSD-NV (1.52%). HC exhibited extreme positive deviations in 1.51% of area, 1.61% of thickness, and 1.18% of volume regions on average. SSD-NV and HoV had smaller percentages of positive deviations, except for HoV in the subcortical volumes (1.72%)(Supplemental table 2 and 3).

**Figure 1.**
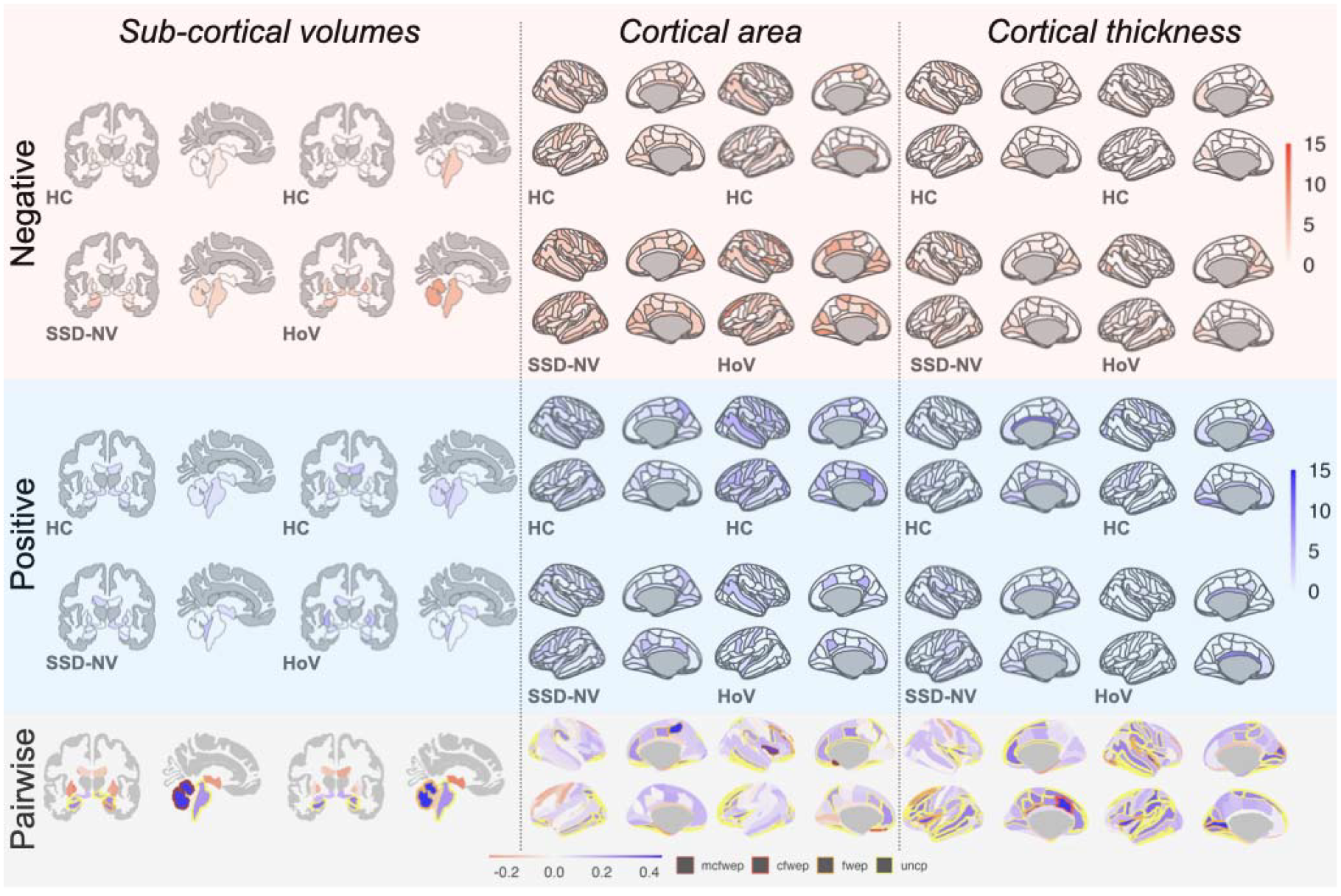
Extreme negative (top) and positive (middle) deviations of subcortical volume (left), cortical area (center) and thickness (right). Color fill represents percentage of participants from total in the group in given ROI. Diagnostic groups (DX) are compared against age matched HC with ratio 1:1. Extreme deviation is defined as |*Z*|>2. (Bottom) pane presents Cohen’s *d* of group differences on the deviation scores. ROIs with significant results are marked with contour lines: *yellow* - uncorrected, *orange* - FDR corrected for the number of ROIs, *red* - FDR corrected for the number of ROIs and number of contrasts and *dark red* - p-value FDR corrected for number of ROIs, contrast and modalities.

HoV and SSD-NV groups showed a significantly higher frequency of negative deviations compared to HC (*p*=.020, *d*=.31, and *p*=.019, *d=*.*48*, respectively). The HoV group also demonstrated a higher frequency of positive deviations (*p*=.009, *d*=.49) Supplemental table 4). No significant differences in the number of positive or negative deviations were observed between HoV and SSD-NV (Supplemental table 4). SSD-NV and HoV had a larger proportion of individuals with extreme deviations across the entire range, compared to the HC (Figure 2). Supplemental figure 4 provides the same analysis with the HoV group stratified SSD-V and nonSSD-V, where the overall pattern of deviations was similar as for the combined group.

**Figure 2.**
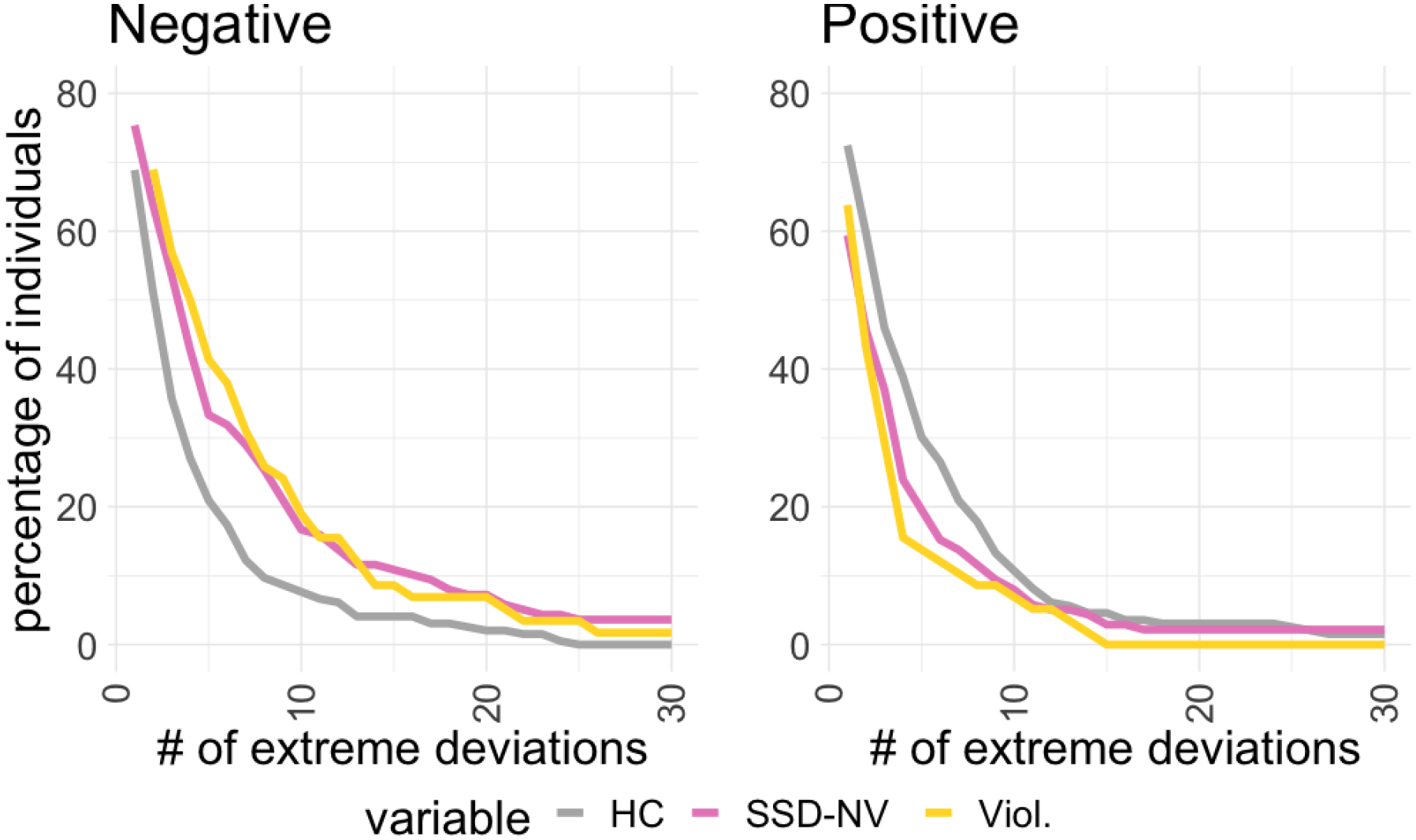
Association between the percentage of individuals as and the frequency of extreme negative (left) or positive (right) deviations in diagnostic categories. Compared to healthy controls (HC), diagnostic groups exhibit a higher prevalence of negative deviations and lower of positive.

### Regional patterns of deviations

The mean of the z-score values of negative deviations were significantly higher (i.e. reflecting stronger or more negative deviations) in the area of the right subcallosal gyrus (*p*_*mc-fwe*_=0.0127, *t*=4.42, *Cohen’s d*=0.98) and superior circular sulcus of the insula (*p*_*mc-fwe*_=0.0312, *t*=4.20, *d*=0.93) in HoV compared to HC. SSD-NV had stronger or more negative deviations in the right (*p*_*mc-fwe*_=0.0150, *t*=4.39, *d*=0.56) and left (*p*_*mc-fwe*_=0.0181, *t*=4.34, *d*=0.55) volume of cerebellum cortex as compared to HC. Among the 332 regions tested across 4 contrasts (1328 tests in total) these regions remained significant after multiple comparison adjustment (Figure 3).

**Figure 3.**
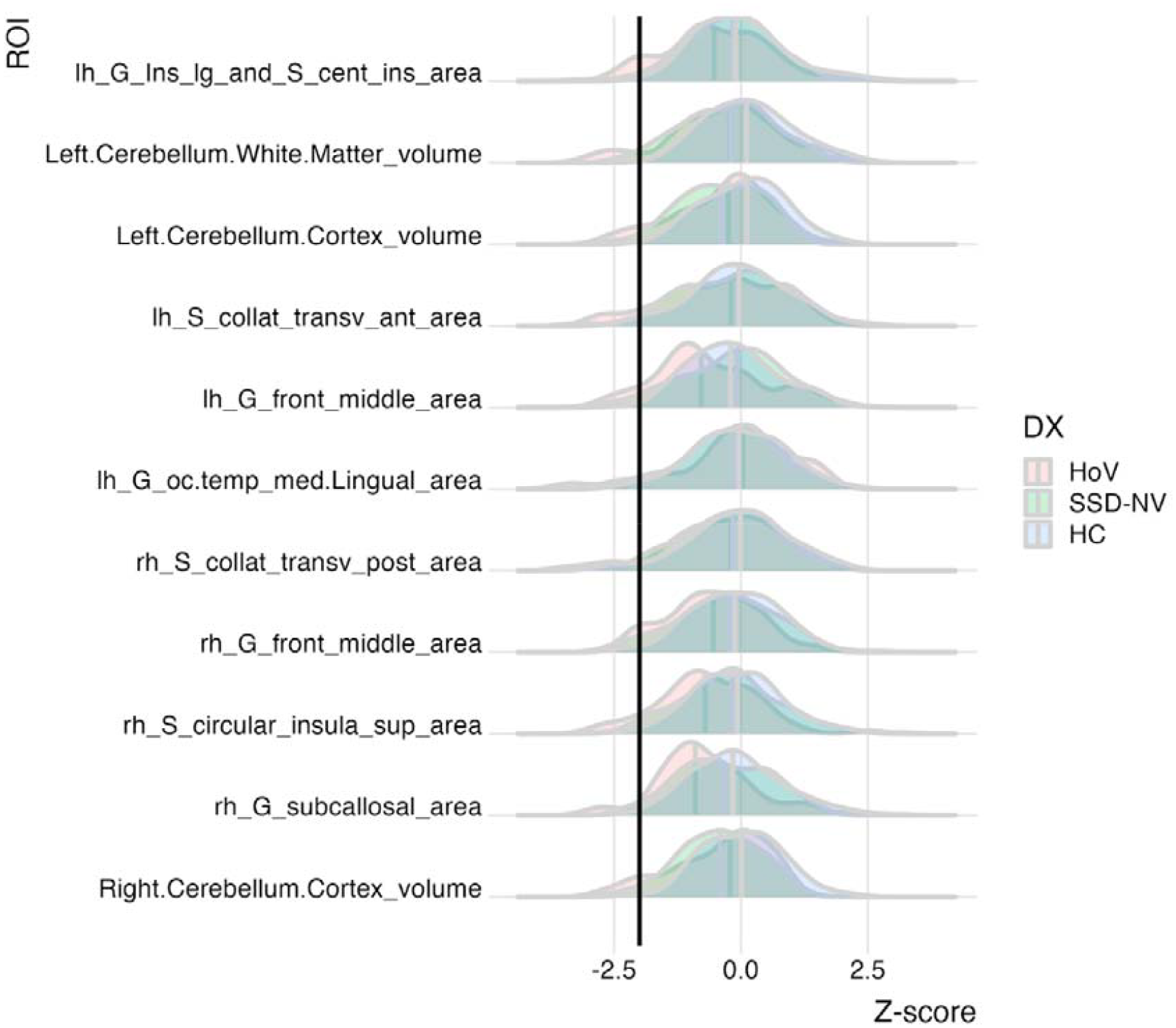
Z-scores indicating deviations from the normative curve for regions with significant group differences are presented. The threshold denoting extremely negative deviations is represented by a black vertical line (Z < -2). It is important to note that mean differences often fail to capture the full range of variations, particularly at the extreme ends.

In the in HoV group, the regions with the largest percentage of individuals with extreme negative deviations were the right and left middle frontal gyrus area (8.62%), right superior circular sulcus of the insula area (8.62%) and right posterior and left anterior transverse collateral sulcus area (8.62%), the right cerebellum cortex (8.62%), and left cerebellar white matter volume (8.62%) (Supplemental table 5, Figure 3). Among the SSD-NV group the regions with the largest percentage of individuals with extreme negative deviations were the left vessel volume (9.42%), right suborbital sulcus area (7.25%) and right the superior occipital and transverse occipital sulcus area (7.25%) (Supplemental table 6, Figure 3).

### Associations with psychopathy traits

PCL-R scores did not significantly differ between HoV participants with and without psychosis (*t*=0.505, *p*=0.45). We found no significant associations between psychopathy scores and brain morphology deviations, but an overall heterogeneous pattern with predominantly positive associations, which was trend level (uncorrected) significant in the right subparietal sulcus area (=1.8, *p*_*unc*_=0.0041) and parieto-inferior supramarginal gyral thickness (*d*=1.8, *p*_*unc*_=0.0056) (Figure 4, Supplemental figure 5). Full list of the associations for each modality is provided in the supplemental table 7.

**Figure 4.**
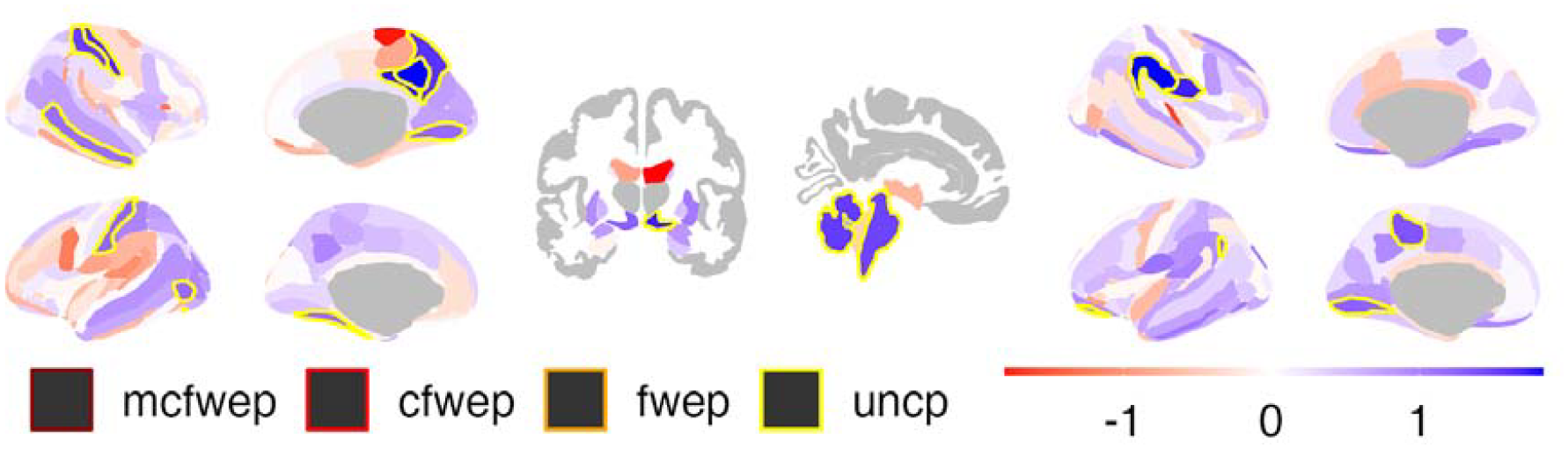
Associations between deviations from norm and psychopathy traits (PCL-R). ROIs fill color represents effect size (Cohen’s *d*) of association. While ROIs contour represents significance of the association: *yellow* - uncorrected, *orange* - FDR corrected for the number of ROIs, *red* - FDR corrected for the number of ROIs and number of contrasts and *dark red* - p-value FDR corrected for number of ROIs, contrast and modalities. None of the results remained significant after adjustment for multiple comparisons, but the overall, heterogeneous PCL-R association pattern is positive (in blue).

## Discussion

In the present study, we used brain imaging and normative modeling to map individual brain morphological deviations among patients with a history of violence and psychosis. The main findings were (1) an overall heterogeneous pattern of deviations on the individual level, with a (2) significantly higher number of extreme negative deviations in persons with a history of violence with or without a schizophrenia spectrum disorder (HoV) and persons with a schizophrenia spectrum disorder but no history of violence (SSD-NV), than healthy controls (HC). (3) These differences were mostly present in cortical area and subcortical volumes, but not in cortical thickness, and (4) significant for regions within the subcallosal and insular cortices and the cerebellum. The study demonstrates the potential of normative modeling to assess brain morphological heterogeneity in forensic psychiatry neuroimaging studies.

Brain based biomarkers of violence proneness in psychosis and psychopathy have long been searched for, with diverging results and limited success (Fjellvang, Groning, et al., 2018; Johanson et al., 2019). Here, we show heterogeneous patterns of brain deviations associated with violence and schizophrenia spectrum disorders. The patterns are highly overlapping (Figure 1), but with differences in subcortical volumes where HoV showed a higher percentage of extreme negative deviations compared to both HC and the SSD-NV group. Moreover, HoV and SSD-NV showed significantly more extreme negative deviations than HC (Figure 2, supplemental table 4) which is in line with previous studies applying normative models to study brain heterogeneity at the individual level in schizophrenia (Wolfers et al., 2018), first episode psychosis (Remiszewski et al., 2022), and conduct disorder (Holz et al., 2022). The regional pattern of extreme deviations does not align with our hypotheses and the results from previous case-control studies where group level volume reductions in amygdala, orbitofrontal cortex, and anterior cingulate have been associated with violence and psychosis ((Fjellvang, Groning, et al., 2018) and (Johanson et al., 2019) for overview). Rather, we found a larger number of negative deviations associated with violence in regions within the middle-frontal gyrus, insula, collateral sulcus, and cerebellum (Table 5). These results comprise cortical regions such as the dorsolateral prefrontal cortex involved in inhibitory control (dorsolateral prefrontal cortex) in schizophrenia patients with a history of violence (Tikàsz et al., 2018) and men with pedophilic disorder (Szczypiński et al., 2022), and the insula known to be involved in empathy, social cognition, and risky decision making (Uddin, Nomi, Hébert-Seropian, Ghaziri, & Boucher, 2017). The cerebellum is important to motor control, cognition, and executive control (D’Angelo, 2018), but has also been linked to aggressive behavior (Leutgeb et al., 2016; Wolfs, Klaus, & Schutter, 2023). Although not overlapping with results from earlier case control-studies, the extreme deviations are found in regions and networks relevant to violent and aggressive behavior.

We found that the patterns of extreme deviations and mean deviations across groups were partly overlapping. With stringent adjustment for multiple comparisons, HoV on average showed more negative deviations in the right superior circular sulcus of the insula (*d*=0.93) compared to HC, a region where HoV showed the largest number of extreme deviations. In addition, HoV showed significantly more negative deviations in the right subcallosal gyrus area (*d*=0.98). The subcallosal gyrus is located adjacent to the anterior cingulate and has been associated with empathy (Rankin et al., 2006) and depression (Dunlop et al., 2023; Sobstyl, Kupryjaniuk, Prokopienko, & Rylski, 2022). Interestingly, the subcallosal gyrus is by some considered as a part of the anterior cingulate (Hamani et al., 2011), which is part of a neural network involved in aggression with e.g. the insula, amygdala and orbitofrontal cortex (Repple et al., 2017). The partly overlapping results between extreme and mean deviations suggest that the patterns of heterogeneity might reflect inter-individual deviations at the extreme end of the normative curve but also with-in and across the normal distribution (Figure 3).

Violence is a heterogenous construct comprising different behaviors, psychological profiles, situational characteristics, and sometimes mental disorders (Whiting et al., 2022) and psychopathy traits (De Brito et al., 2021). We found no associations between psychopathy scores and deviation patterns, but a positive trend (significance at the uncorrected level) in the supramarginal gyrus area, which is involved in emotion processing (Silani, Lamm, Ruff, & Singer, 2013). The HoV group consisted of persons with and without schizophrenia with similar levels of psychopathy traits, and the patterns of deviances were different between HoV and SSD-NV. Indeed, the largest study to date comparing psychopathy and brain morphology in violent offenders with and without schizophrenia report group differences in specific regions that do not overlap with the regions where we observe extreme or greater average deviations in HoV (Kolla et al., 2021). Taken together, this suggests the deviations might reflect the violence proneness independent of psychosis status as a trans-diagnostic feature. Comparatively, a recent study assessing aggression in a group of adolescents with diagnoses across the oppositional - conduct disorder spectrum merged together, showed inter-individual aggression related deviation patterns (Holz et al., 2022).

The study has some limitations. The number of subjects in the HoV group was limited, although similar to previous case-control imaging studies investigating violence in schizophrenia and psychopathy (e.g. (Del Bene et al., 2016; Kolla et al., 2021)), and normative modeling of aggression in adolescents (Holz et al., 2022). In normative modeling, as in regular case-control studies, the variance within volume estimates tends to decrease as sample size increases, but this decrease is modest (Bozek, Griffanti, Lau, & Jenkinson, 2023). The variability bias is not improved much with larger sample sizes and is greatest at the extreme age-range (Bozek et al., 2023) which is not included in the current study. Moreover, although the Freesurfer Destrieux atlas comprises 148 cortical ROIs, atlas defined cortical parcellations have shown lower representations of true associations and prediction accuracy than vertex-wise analyses (Fürtjes, Cole, Couvy-Duchesne, & Ritchie, 2023). However, the normative charts derived from >50 000 individuals are based on this Freesurfer standard atlas (Rutherford et al., 2022) which ensures that the results are reproducible and easy to compare, and facilitates replication studies. An inherent limitation in all cross-sectional neuroimaging studies is the inability to distinguish between specific disease mechanisms and the effects of early-life influences and developmental trajectories. Cross-sectional normative modeling has shown lower individual level prediction, propensity than longitudinal models (Di Biase et al., 2023), which should also be employed in future studies of violence and psychosis. However, it is worth noting that our study encompassed a broad age range providing comprehensive coverage across a significant fraction of the lifespan.

In conclusion, this proof-of-principle study demonstrates the feasibility of normative modeling to map a heterogeneous pattern of brain morphometry deviations associated with violence and psychosis, and extends the regional group difference patterns from classical case-control studies. While the current results warrant replication and further integration with environmental and psychological variables, studies addressing individual brain deviations may contribute to improved understanding of the complex and multidimensional underpinnings of violence in forensic psychiatry.

## Ethical standards

The authors assert that all procedures contributing to this work comply with the ethical standards of the relevant national and institutional committees on human experimentation and with the Helsinki Declaration of 1975, as revised in 2008.

## Data Availability

All data produced in the present study are available upon reasonable request to the authors

