## Supplementary material for "Individual-level deviations from normative brain morphology in violence, psychosis, and psychopathy"

### Supplementary results

When we did not adjust for the number of modalities (i.e. cortical thickness, cortical area or subcortical regions), we also observed more negative deviations in the orbital medial olfactory sulcus area in the HoV group compared to healthy controls (pc-fwe=0.026, pmc-fwe=0.0542, t=4.0629, cD=0.90).

The SSD-NV group showed stronger or more positive deviations in the left inferior lateral ventricle volume (pc-fwe=0.0102, pmc-fwe=0.0822, t=3.95, cD=0.50) and stronger negative deviations in left cerebellum white matter volume (pc-fwe=0.0173, pmc-fwe=0.1519, t=3.76, cD=0.48) compared to healthy controls. Additionally, the SSD-NV group exhibited more negative deviations in thickness of mid-anterior cingulate gyrus on the left hemisphere (pc-fwe=0.0404, pmc-fwe=0.1022, t=3.88, cD=0.47).

### Supplementary figures

##### **Supplemental figure 1.** Extreme negative and positive deviations of cortical area

|  | **nonSSD-V** | **SSD-NV** | **SSD-V** |
| --- | --- | --- | --- |
| **HC**  HC | 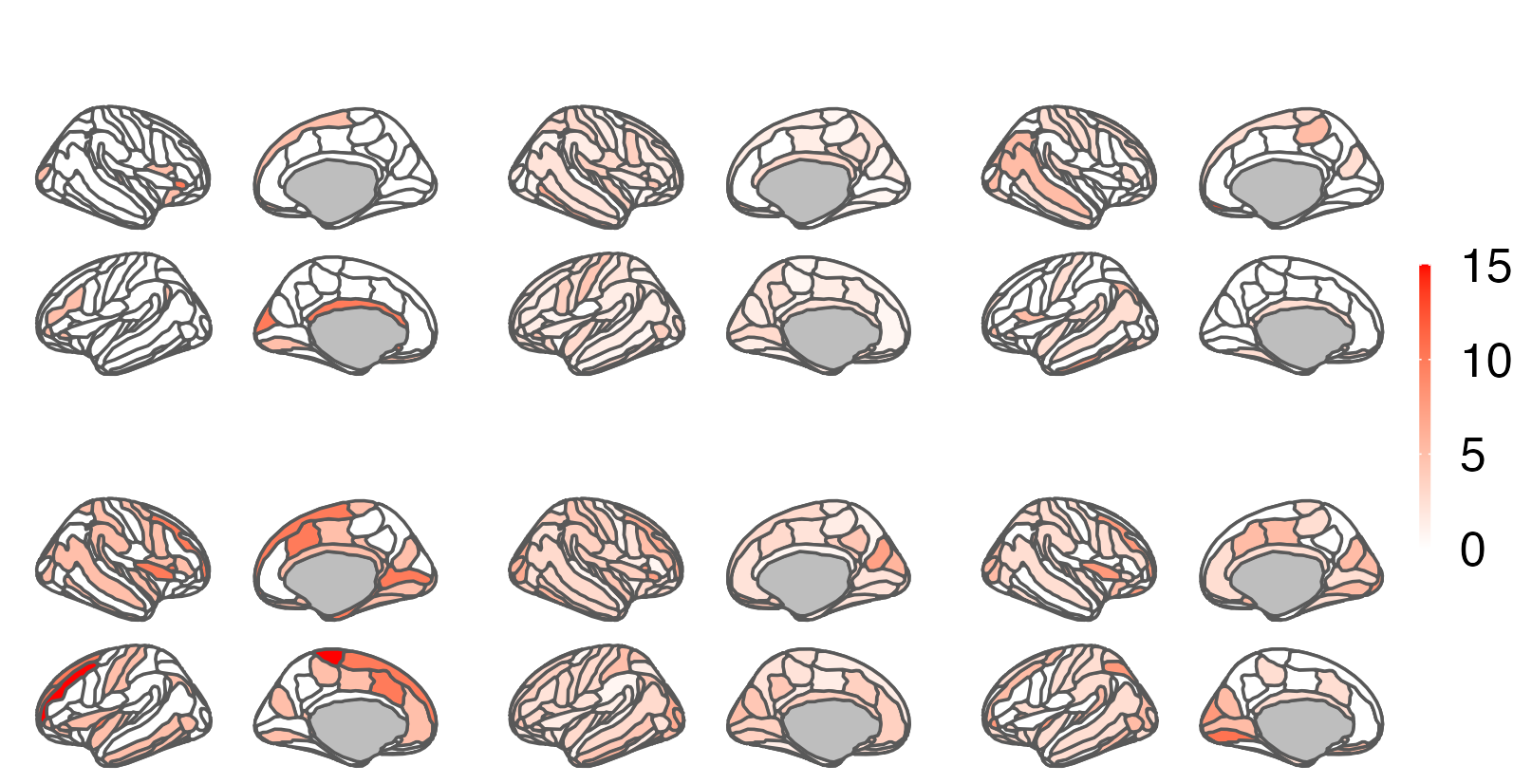 | | |
| **DX**  Diagnostic g. |  |  |  |
| **HC**  HC | 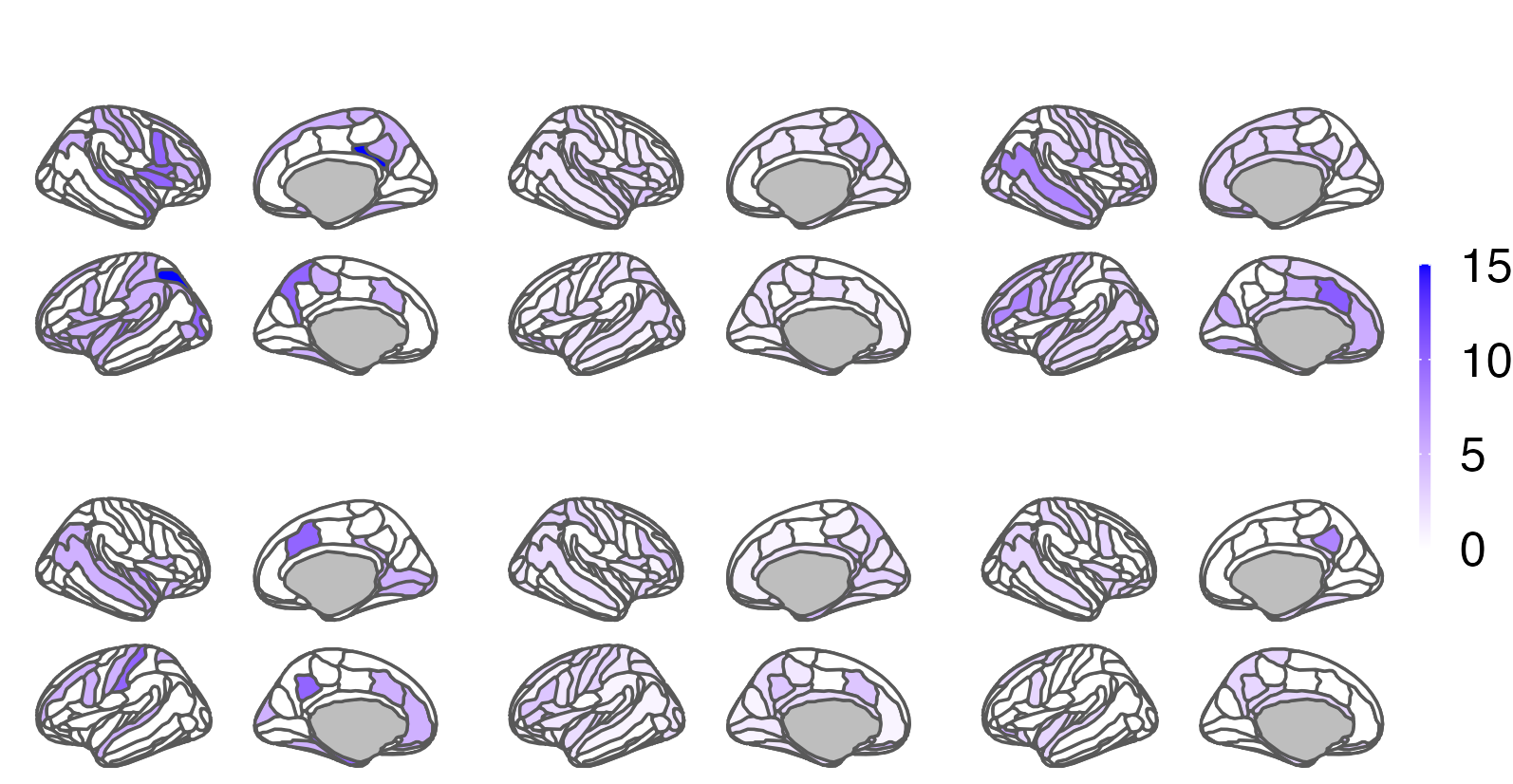 | | |
| **DX**  Diagnostic g. |  |  |  |
| **CD**  Pairwise tests | 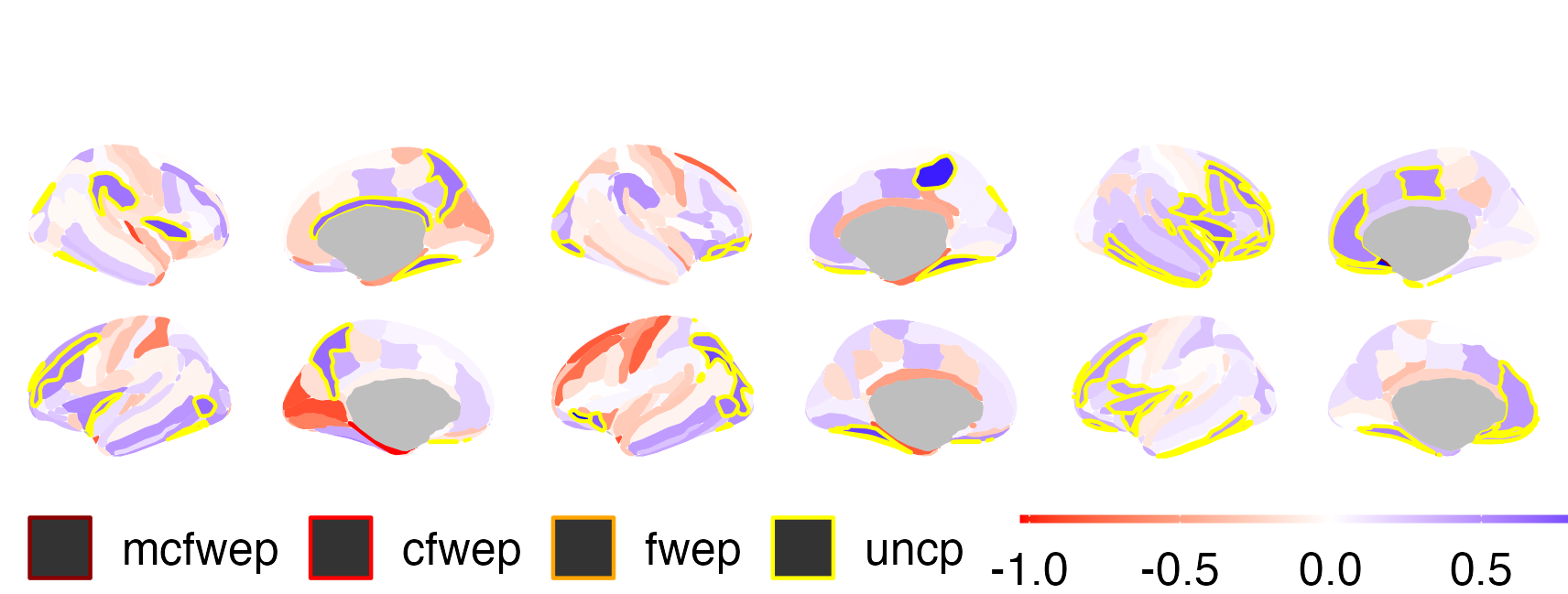 | | |
| Extreme negative (top) and positive (middle) deviations of cortical area. Color fill represents percentage of participants from total in the group in given ROI. Diagnostic groups (DX) are compared against age matched healthy controls (HC) with ratio 1:1. Extreme deviation is defined as \|Z\|>2. (Bottom) pane presents Cohen's *d* (CD) of group differences on the deviation scores. ROIs with significant results are marked with contour lines: *yellow* - uncorrected, *orange* - FDR corrected for the number of ROIs, *red* - FDR corrected for the number of ROIs and number of contrasts and *dark red* - p-value FDR corrected for number of ROIs, contrast and modalities. | | | |

##### **Supplemental figure 2.** Extreme negative and positive deviations of cortical thickness.

|  | **nonSSD-V** | **SSD-NV** | **SSD-V** |
| --- | --- | --- | --- |
| **HC** | 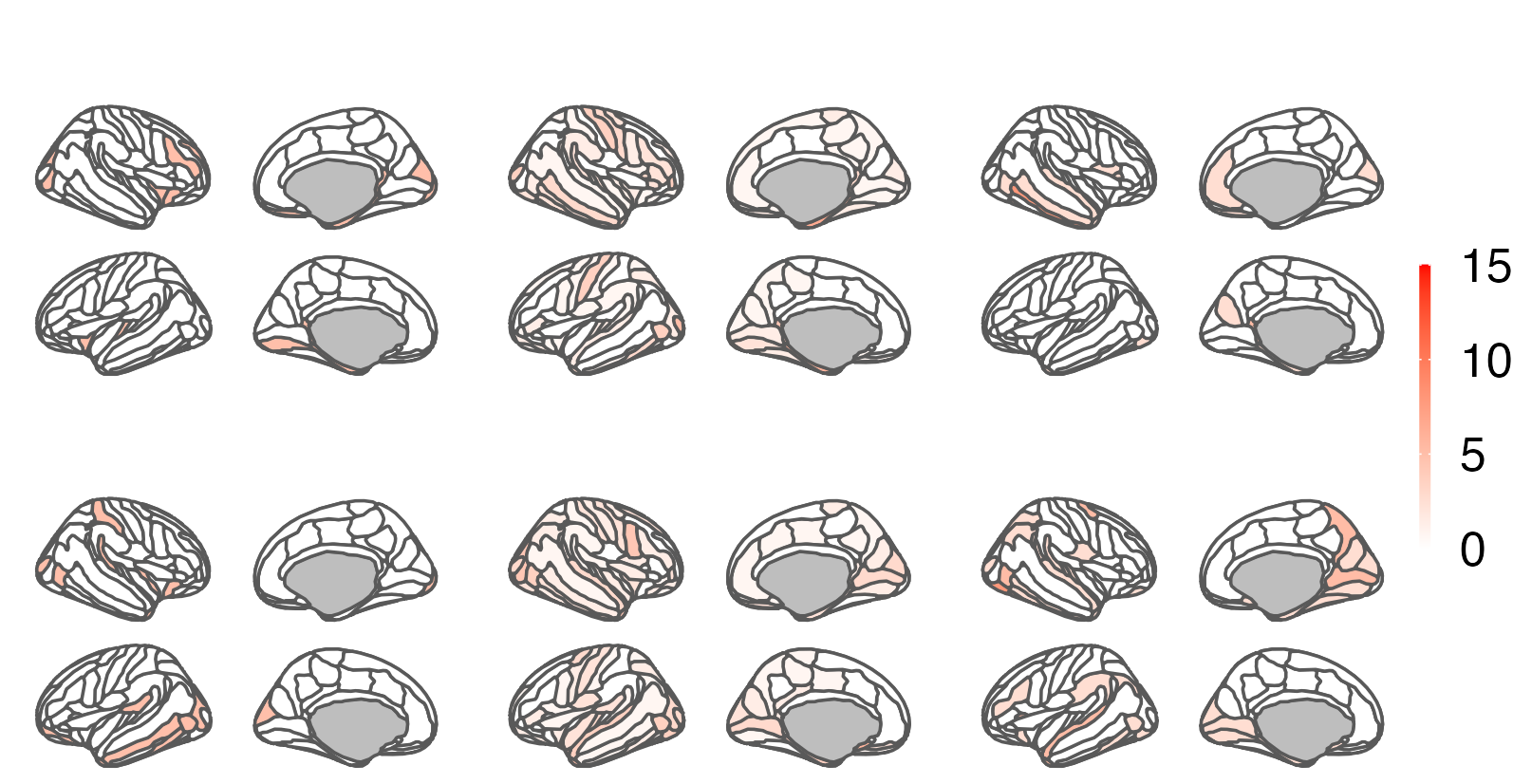 | | |
| **DX** |  |  |  |
| **HC** | 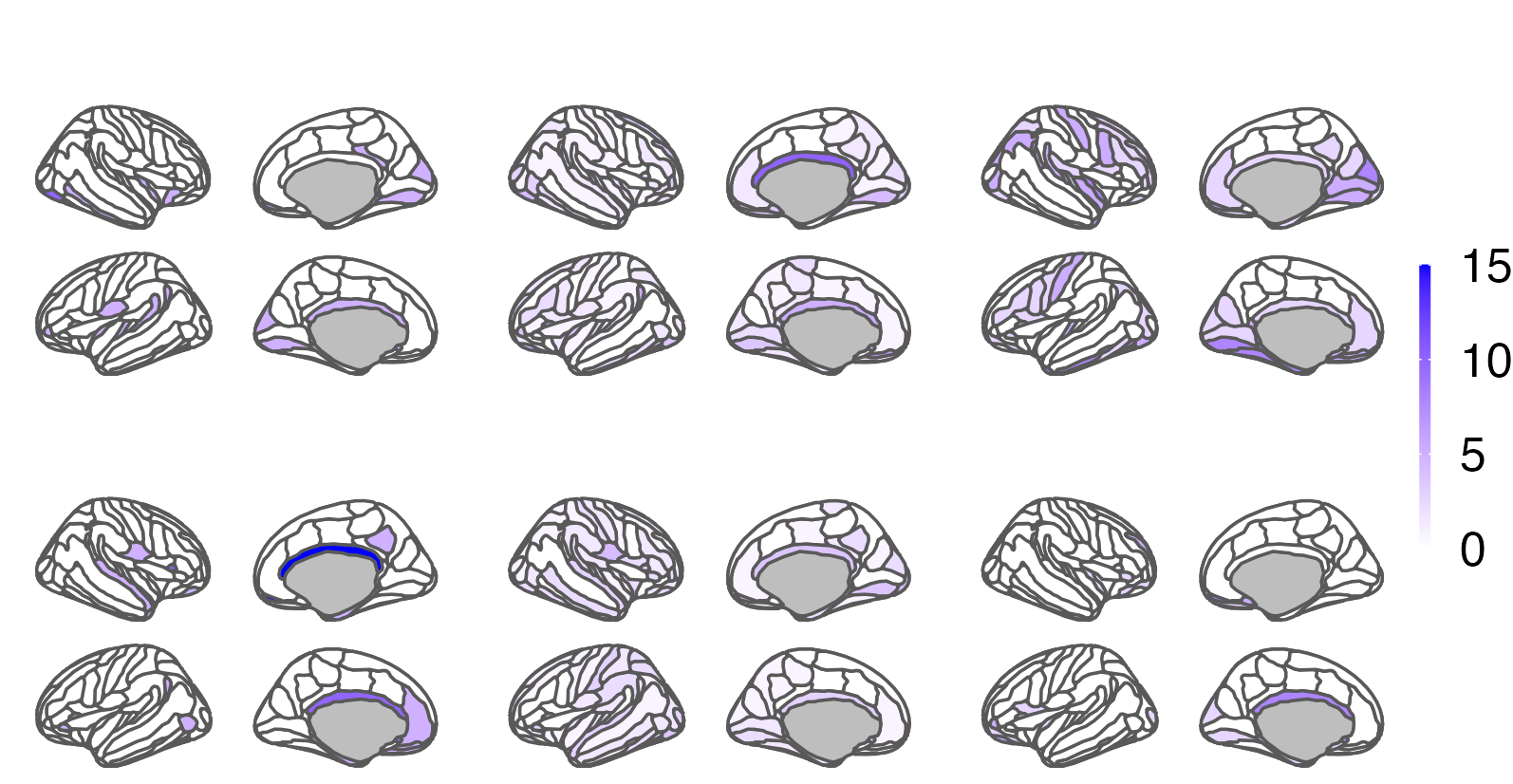 | | |
| **DX** |  |  |  |
| **CD**  Pairwise tests | 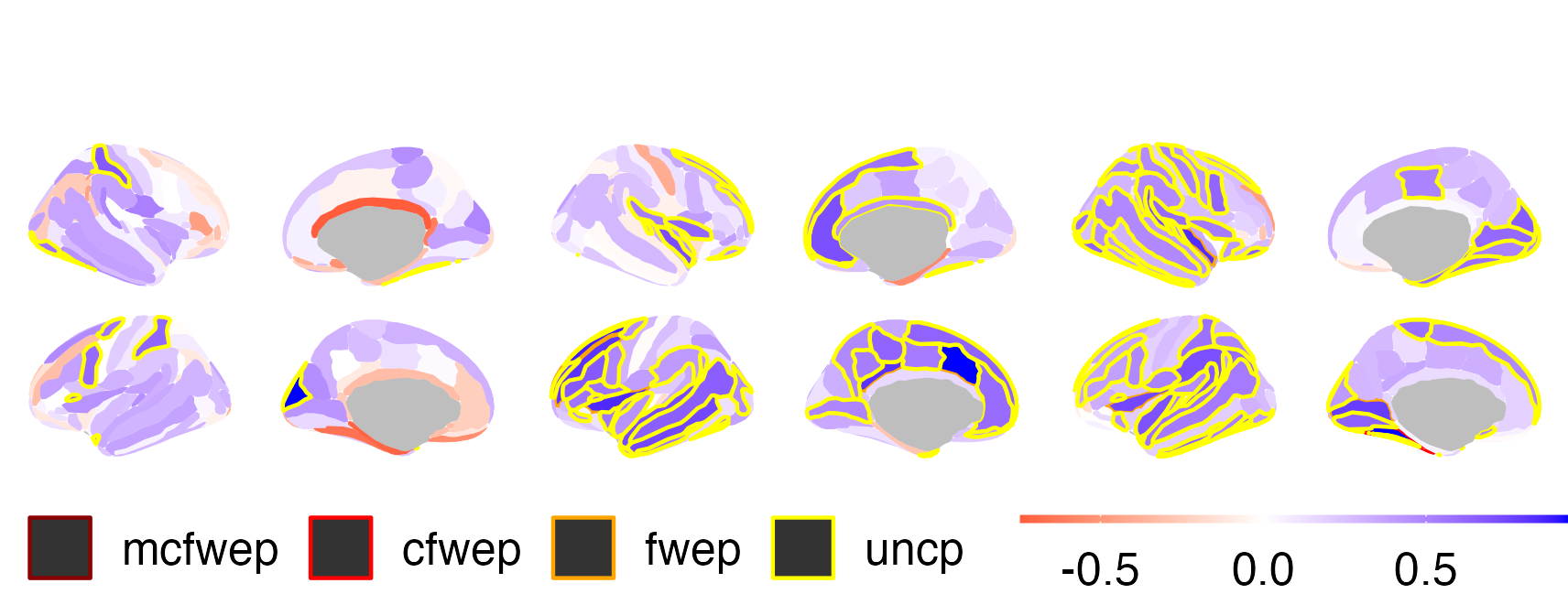 | | |
| Extreme negative (top) and positive (middle) deviations of cortical thickness. Color fill represents percentage of participants from total in the group in given ROI. Diagnostic groups (DX) are compared against age matched healthy controls (HC) with ratio 1:1. Extreme deviation is defined as \|Z\|>2. (Bottom) pane presents Cohen's *d* (CD) of group differences on the deviation scores. ROIs with significant results are marked with contour lines: *yellow* - uncorrected, *orange* - FDR corrected for the number of ROIs, *red* - FDR corrected for the number of ROIs and number of contrasts and *dark red* - p-value FDR corrected for number of ROIs, contrast and modalities. | | | |

##### **Supplemental figure 3.** Extreme positive and negative deviations of subcortical volumes.

|  | **nonSSD-NV** | **SSD-NV** | **SSD-V** |
| --- | --- | --- | --- |
| **HC** | 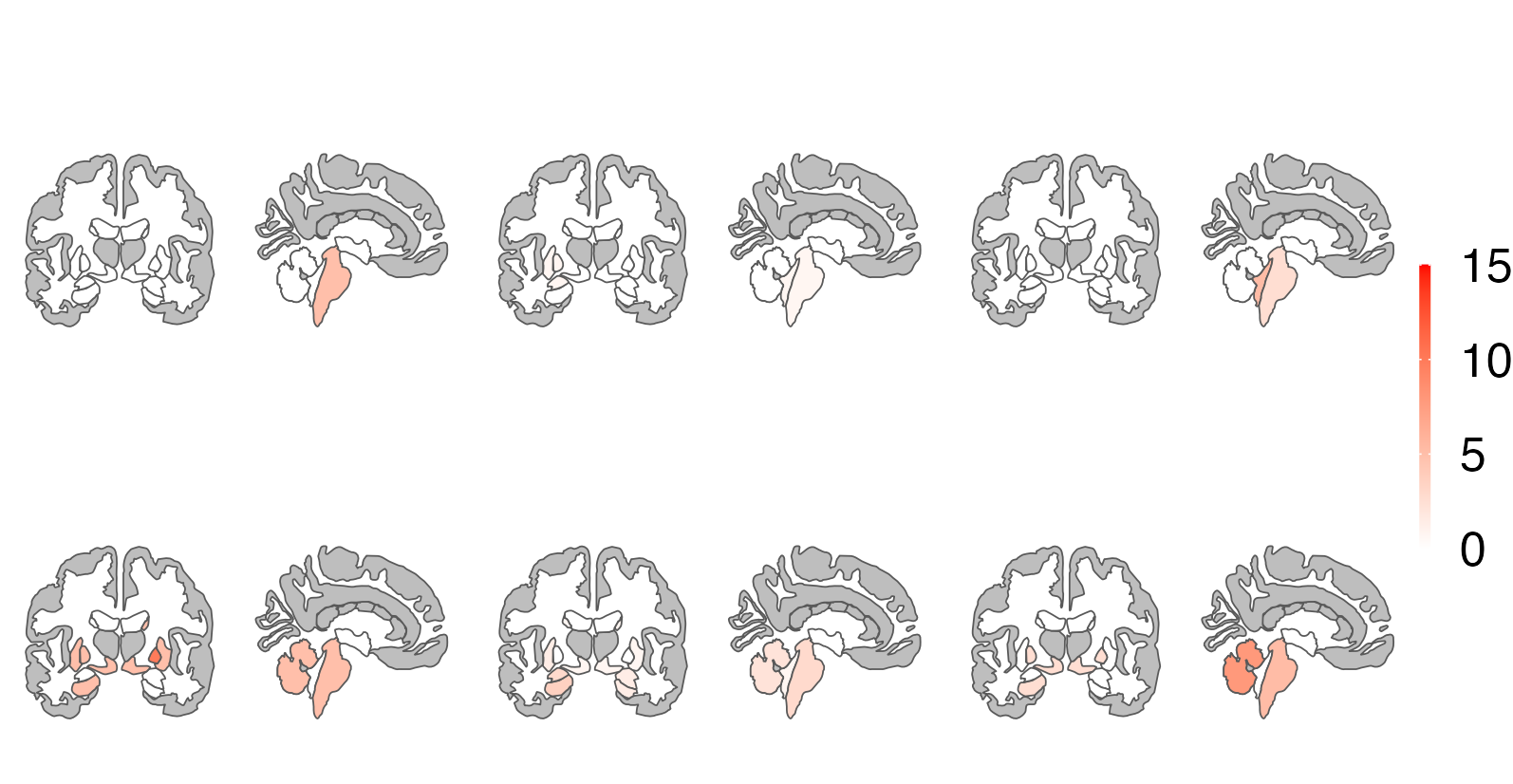 | | |
| **DX** |  |  |  |
| **HC** | 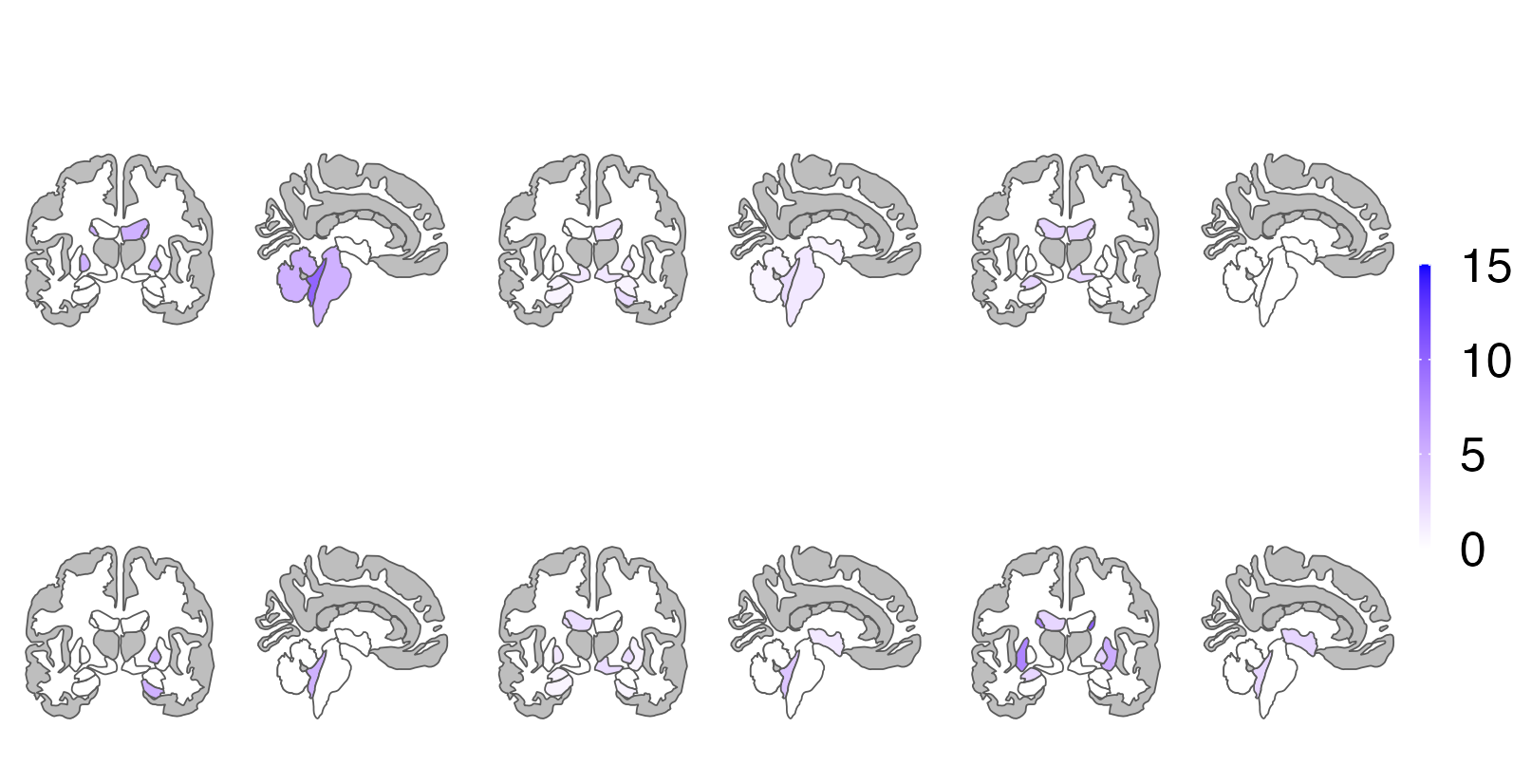 | | |
| **DX** |  |  |  |
| **CD**  Pairwise tests | 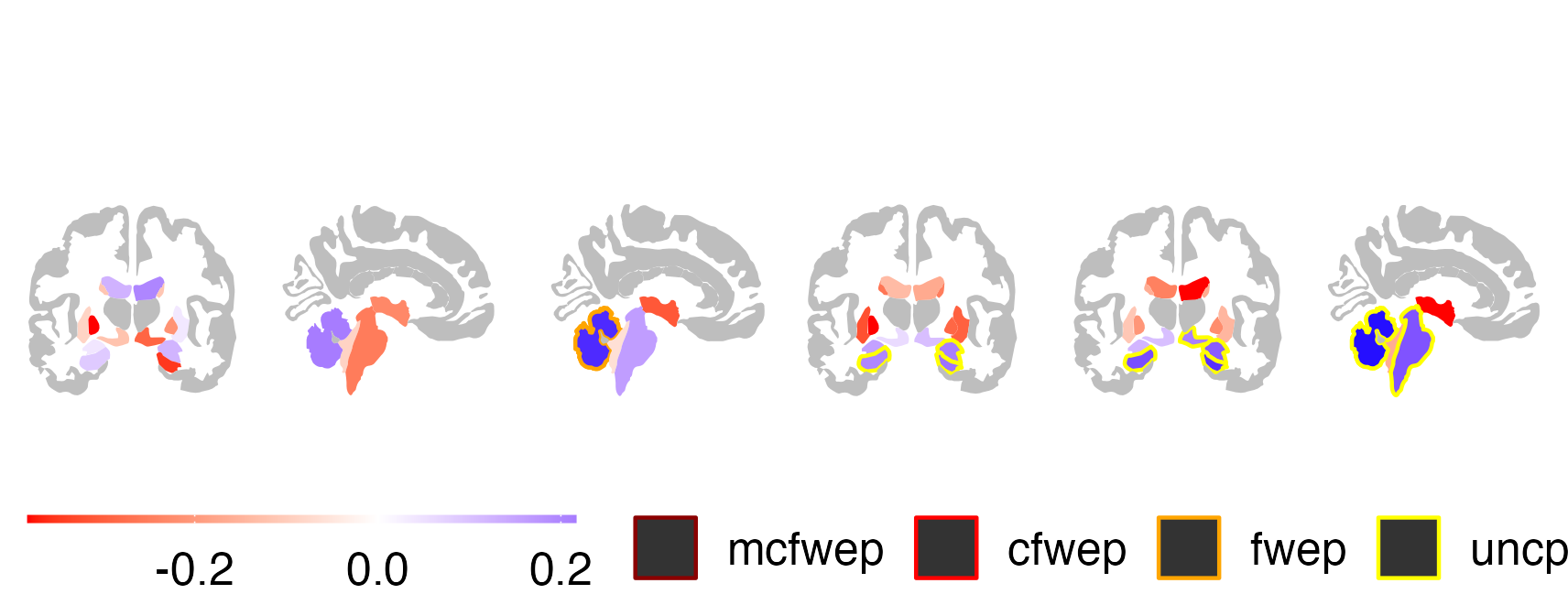 | | |
| Extreme positive (top) and negative (middle) deviations of subcortical volumes. Color fill represents percentage of participants from total in the group in given ROI. Diagnostic groups (DX) are compared against age matched healthy controls (HC) with ratio 1:1. Extreme deviation is defined as \|Z\|>2. (Bottom) pane presents Cohen's *d* (CD) of group differences on the deviation scores. ROIs with significant results are marked with contour lines: *yellow* - uncorrected, *orange* - FDR corrected for the number of ROIs, *red* - FDR corrected for the number of ROIs and number of contrasts and *dark red* - p-value FDR corrected for number of ROIs, contrast, and modalities. | | | |

##### **Supplemental figure 4.** Association between the percentage of individuals as and the frequency of extreme negative or positive deviations in diagnostic categories.

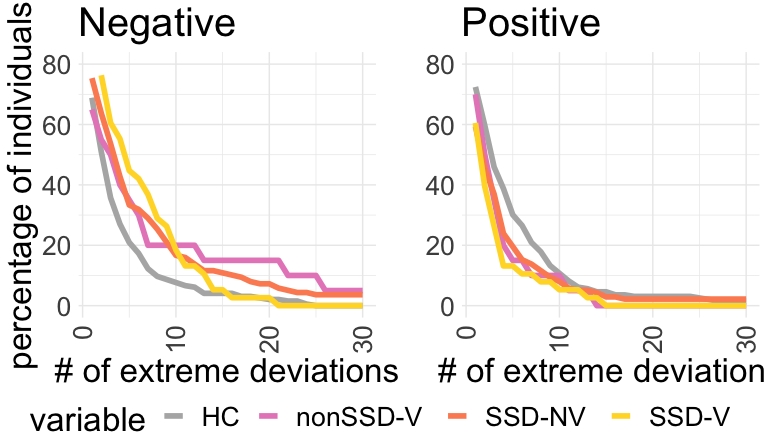

Compared to healthy controls (HC), diagnostic groups exhibit a higher prevalence of negative deviations and lower of positive. Notably, in the SSD-V group, the prevalence of extreme negative deviations approaches that of HC when the number of outliers reaches 15, whereas in the SSD-NV and nonSSD-V groups, it remains elevated.

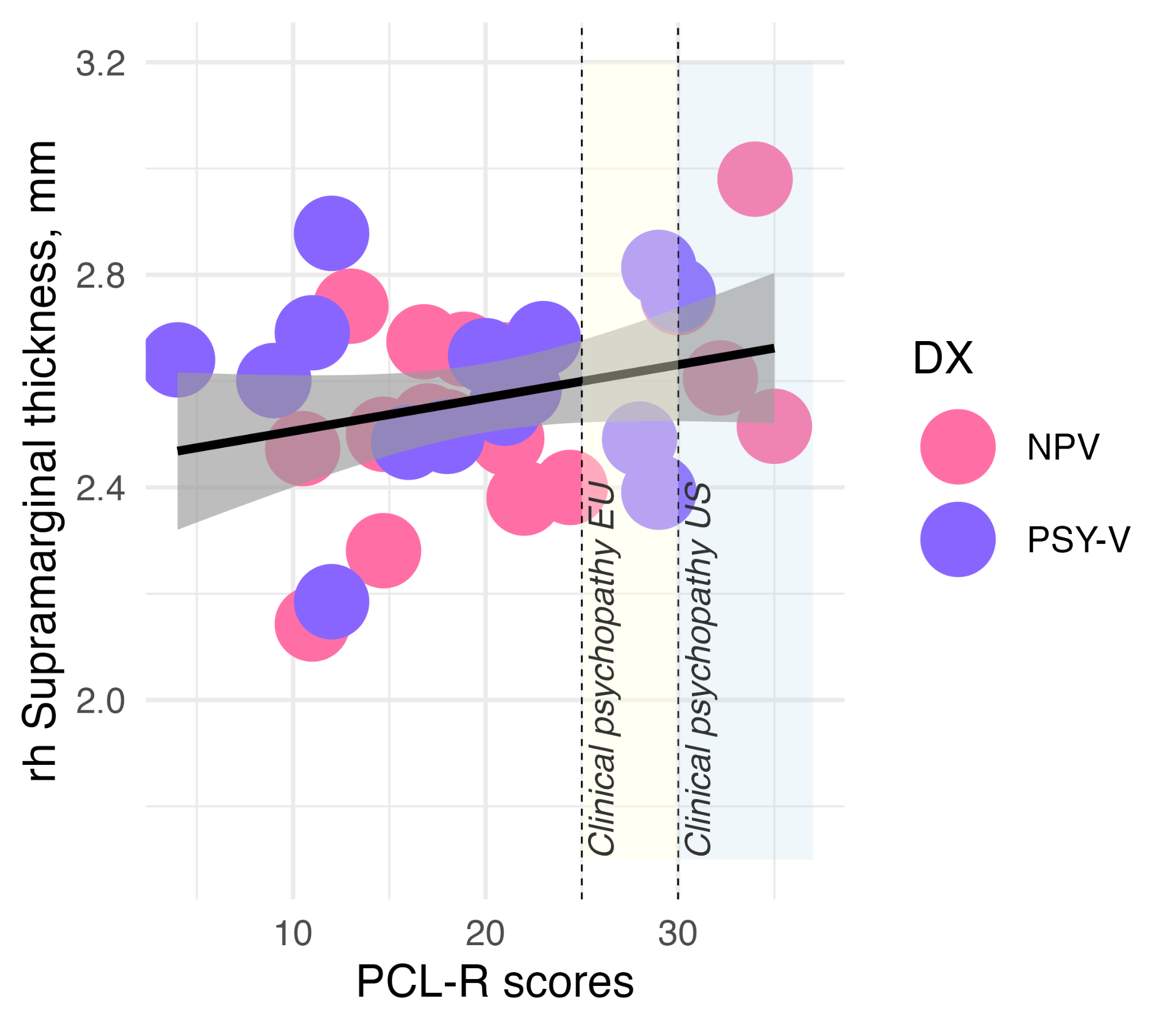

##### **Supplemental figure 5.** Association between psychopathy traits and cortical thickness.

Association between psychopathy traits (PCL-R scores) and cortical thickness in the right hemisphere supramarginal gyrus, among persons with a history of violence with (purple) or without (pink) a comorbid schizophrenia spectrum disorder.

### Supplementary tables

##### **Supplemental table 1.** Demographics and clinical characteristics

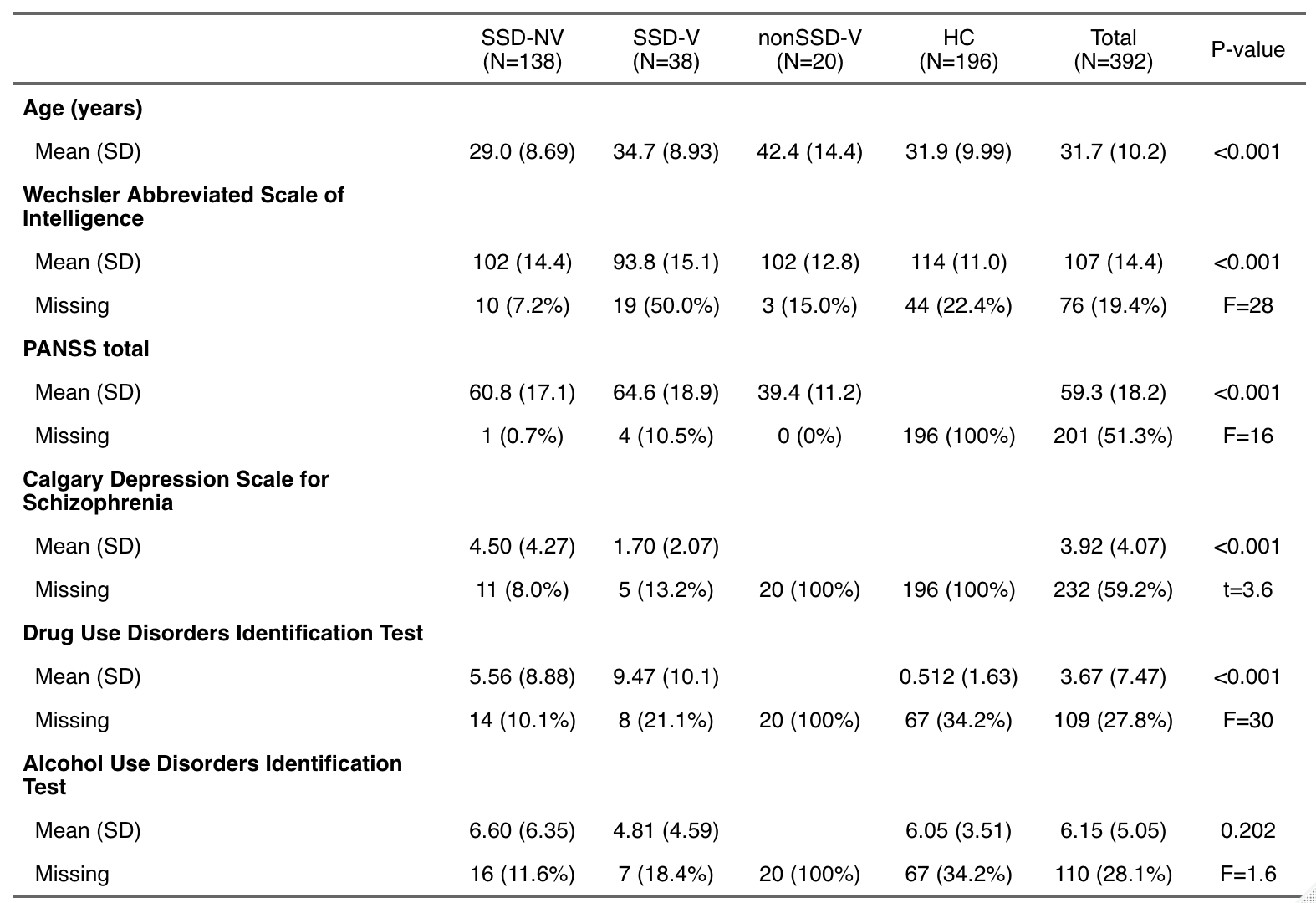

Abbreviations: SSD-NV - schizophrenia spectrum disorder patients without history of violence, SSD-V - schizophrenia spectrum disorder patients with a history of violence, nonSSD-V - participants with history of violence and no schizophrenia spectrum disorder, HC - healthy controls.

##### **Supplemental table 2.** Percentage of extreme negative and positive deviations for each group

|  | **Negative** | | | | **Positive** | | | |
| --- | --- | --- | --- | --- | --- | --- | --- | --- |
| **DX** | **Area**  *n*=150 | **Thickness**  *n*=150 | **Volume**  *n*=32 | **Total**  *n*=332 | **Area**  *n*=150 | **Thickness**  *n*=150 | **Volume**  *n*=32 | **Total**  *n*=332 |
| **HC** | 1.51 | 0.95 | 0.59 | 1.17 | 1.56 | 1.61 | 1.18 | 1.55 |
| **SSD-NV** | 3.07 | 1.33 | 1.52 | 2.13 | 1.23 | 1.30 | 1.11 | 1.25 |
| **HoV** | 2.75 | 1.09 | 2.42 | 1.97 | 0.86 | 0.81 | 1.72 | 0.92 |

For each group, we calculated the percentage of extreme negative and positive deviations, defined as |Z|>2 per modality. The header of each modality indicates the total number of ROIs. Our results showed that diagnostic groups exhibited a higher frequency of negative outliers in area and volume, as compared to healthy controls, but not in cortical thickness. Moreover, diagnostic groups had either fewer or similar instances of extreme positive deviations across all three modalities, with exception of volumes for the HoV group. *Abbreviations:* DX - diagnosis, SSD-NV - schizophrenia spectrum disorder patients without history of violence, HoV - participants with history of violence, HC - healthy controls.

##### **Supplemental table 3.** Percentage of extreme negative and positive deviations

|  | **Negative** | | | | **Positive** | | | |
| --- | --- | --- | --- | --- | --- | --- | --- | --- |
| **DX** | **Area**  *n*=150 | **Thick.**  *n*=150 | **Volume**  *n*=32 | **Total**  *n*=332 | **Area**  *n*=150 | **Thickness**  *n*=150 | **Volume**  *n*=32 | **Total**  *n*=332 |
| **HC** | 1.51 | 0.95 | 0.59 | 1.17 | 1.56 | 1.61 | 1.18 | 1.55 |
| **nonSSD-V** | 3.07 | 0.93 | 2.97 | 2.09 | 1.20 | 0.93 | 0.94 | 1.05 |
| **SSD-NV** | 3.07 | 1.33 | 1.52 | 2.13 | 1.23 | 1.30 | 1.11 | 1.25 |
| **SSD-V** | 2.58 | 1.18 | 2.14 | 1.90 | 0.68 | 0.75 | 2.14 | 0.86 |

For each group, we calculated the percentage of extreme negative and positive deviations, defined as |Z|>2, for each ROI. The header of each modality indicates the total number of ROIs. Our results showed that all three diagnostic groups exhibited a higher frequency of negative outliers in area and volume, as compared to healthy controls, but not in cortical thickness. Moreover, these diagnostic groups had either fewer or similar instances of extreme positive deviations across all three modalities. *Abbreviations:* DX - diagnosis, SSD-NV - schizophrenia spectrum disorder patients without history of violence, SSD-V - schizophrenia spectrum disorder patients with a history of violence, nonSSD-V - participants with history of violence and no schizophrenia spectrum disorder, HC - healthy controls.

##### **Supplemental table 4.** Pairwise comparisons of frequency of extreme deviations between clinical groups in individuals

| **contrast** | ***p*_uncorrected_** | ***p*_c-fwe_** | **Cohen’s *d*** |
| --- | --- | --- | --- |
|  | **Extreme negative deviations** | | |
| HC>SSD-NV | **0.0034** | **0.0195** | 0.3148 |
| HC>HoV | **0.0067** | **0.0191** | 0.4782 |
| SSD-NV>HoV | 0.3419 | 0.7213 | 0.0664 |
|  | **Extreme positive deviations** | | |
| HC>SSD-NV | 0.2895 | 0.6227 | 0.0731 |
| HC>HoV | **0.0053** | **0.0093** | 0.4882 |
| SSD-NV>HoV | 0.1637 | 0.425 | 0.2102 |

Pairwise comparisons of frequency of extreme deviations between clinical groups in individuals, significant results are marked in bold. *Abbreviations:* SSD-NV - schizophrenia spectrum disorder patients without history of violence, HoV - participants with history of violence, HC - healthy controls.

##### **Supplemental table 5.** Regions with the highest percentage of individuals with extreme negative deviations

| **Hemisphere** | **Region** | **Modality** | **SSD-NV** | **HoV** | **HC** |
| --- | --- | --- | --- | --- | --- |
| Left | Vessel | volume | 9.42 | 3.45 | 4.59 |
| Right | S_suborbital | area | 7.25 | 1.72 | 6.63 |
| Right | S_oc_sup_and_transversal | area | 7.25 | 3.45 | 3.06 |
| Right | Lat_Fis.ant.Vertical | area | 7.25 | 5.17 | 2.04 |
| Right | Lat_Fis.ant.Horizont | area | 7.25 | 1.72 | 4.08 |
| Right | S_parieto_occipital | area | 7.25 | 5.17 | 1.02 |
| Right | G_occipital_middle | area | 6.52 | 5.17 | 1.02 |
| Left | S_oc_middle_and_Lunatus | area | 6.52 | 1.72 | 2.04 |
| Right | G_front_middle | area | 6.52 | 8.62 | 2.04 |
| Left | G_occipital_middle | area | 6.52 | 3.45 | 1.02 |
| Left | G_subcallosal | thickness | 5.80 | 1.72 | 0.51 |
| Left | S_oc_sup_and_transversal | area | 5.80 | 0.00 | 4.59 |
| Left | S_temporal_inf | area | 5.80 | 1.72 | 3.57 |

Regions with the highest percentage of individuals with extreme negative deviations, focusing exclusively on diagnostic groups. The table is arranged in descending order based on percentages in the SSD-NV column. *Abbreviations:* SSD-NV - schizophrenia spectrum disorder patients without history of violence, HoV - participants with history of violence, HC - healthy controls.

##### **Supplemental table 6.** Regions with the highest percentage of individuals with extreme negative deviations

| **Hemisphere** | **Region** | **Modality** | **SSD-NV** | **HoV** | **HC** |
| --- | --- | --- | --- | --- | --- |
| Right | G_front_middle | area | 6.52 | 8.62 | 2.04 |
| Right | S_circular_insula_sup | area | 3.62 | 8.62 | 1.02 |
| Right | S_collat_transv_post | area | 3.62 | 8.62 | 4.59 |
| Left | G_oc.temp_med.Lingual | area | 3.62 | 8.62 | 1.53 |
| Left | G_front_middle | area | 3.62 | 8.62 | 1.02 |
| Right | Cerebellum.Cortex | volume | 2.90 | 8.62 | 1.02 |
| Left | S_collat_transv_ant | area | 2.90 | 8.62 | 2.04 |
| Left | Cerebellum.White.Matter | volume | 2.17 | 8.62 | 0.00 |
| Left | G_Ins_lg_and_S_cent_ins | area | 2.17 | 8.62 | 0.00 |
| Left | S_orbital_lateral | area | 4.35 | 6.90 | 2.55 |
| Right | Pole_occipital | area | 2.90 | 6.90 | 0.51 |
| Right | G_and_S_cingul.Mid.Ant | area | 2.90 | 6.90 | 0.00 |
| Right | S_collat_transv_ant | area | 2.90 | 6.90 | 2.04 |

Regions with the highest percentage of individuals with extreme negative deviations, stratified by diagnostic groups. The table is arranged in descending order based on percentages in the HoV column. *Abbreviations:* SSD-NV - schizophrenia spectrum disorder patients without history of violence, HoV - participants with history of violence, HC - healthy controls.

##### **Supplemental table 7.** Associations between PCL-R scores and deviations values.

| Feature | hemisphere | modality | direction | cD | T test | p_unc_ | p_fwe_ | p_cfwe_ | p_mcfwe_ |
| --- | --- | --- | --- | --- | --- | --- | --- | --- | --- |
| S_subparietal | right | area | Inc | 1.836 | 2.9489 | 0.0041 | 0.3095 | 0.6922 | 0.8916 |
| G_and_S_paracentral | right | area | Dec | 1.8012 | 2.8930 | 0.0054 | 0.3602 | 0.7325 | 0.9136 |
| G_cingul.Post.dorsal | right | area | Inc | 1.7743 | 2.8498 | 0.0051 | 0.3663 | 0.7633 | 0.9302 |
| Lateral.Ventricle | right | volume | Dec | 1.4417 | 2.3155 | 0.0158 | 0.2811 | 0.6065 | 0.9988 |
| VentralDC | right | volume | Inc | 1.3595 | 2.1835 | 0.0216 | 0.3388 | 0.7019 | 0.9997 |
| Cerebellum.Cortex | left | volume | Inc | 1.2099 | 1.9433 | 0.0349 | 0.4797 | 0.8475 | 1 |
| G_pariet_inf.Supramar | right | thickness | Inc | 1.7759 | 2.7588 | 0.0056 | 0.3748 | 0.7599 | 0.9567 |
| G_and_S_subcentral | right | thickness | Inc | 1.6644 | 2.5857 | 0.0076 | 0.4705 | 0.8689 | 0.9861 |
| G_temp_sup.G_T_transv | right | thickness | Dec | 1.5715 | 2.4413 | 0.0103 | 0.5452 | 0.9360 | 0.9953 |

Associations between PCL-R scores and deviations values. The top three associations for each modality (area, thickness, and volume), as determined by Cohen's d effect size (cD), are presented.
